# Resting-State Functional Connectivity of the Fronto-Limbic and Default Mode Networks as Predictors of Antidepressant Response in Major Depressive Disorder

**DOI:** 10.1101/2025.08.26.25334436

**Authors:** Youbin Kang, JeYoung Jung, Kyu-Man Han, Dorothee P. Auer, Byung-Joo Ham

## Abstract

**Background:** Major depressive disorder (MDD) is a leading cause of disability worldwide, yet treatment response to antidepressants remains highly variable, with a significant proportion of patients failing to achieve remission. Resting-state functional connectivity (RSFC) studies have linked MDD to large-scale brain network dysregulation, but methodological inconsistencies and limited focus on treatment response have hindered clinical translation.

**Methods:** In this study, we conducted a resting-state fMRI study as part of an 8-week antidepressant trial on 86 patients with MDD and 93 healthy controls (HCs). Utilizing a seed-to-voxel approach with brain regions identified in previous studies on neural networks implicated in antidepressant response, we examined RSFC patterns associated with treatment outcomes. Additionally, we explored their relationships with cognitive function and depressive severity to elucidate the neural mechanisms underlying treatment response.

**Results:** Compared to the non-responder group, antidepressant responders exhibited significantly higher FCs between the prefrontal cortex, thalamus, amygdala, and posterior cingulate cortex (PCC). In responders, FC between the left amygdala and supramarginal gyrus, as well as the ventromedial prefrontal cortex (VMPFC) and lateral occipital cortex, was negatively correlated with cognitive scores. Conversely, in the non-responder group, FC between the left subgenual anterior cingulate cortex (sgACC) and lateral occipital cortex was negatively correlated with digit span but positively with other cognitive tasks. FCs involving the amygdala, thalamus, and VMPFC were correlated with depression scale scores.

**Conclusions:** The present study highlights the role of intrinsic FC patterns in differentiating antidepressant responders from non-responders in MDD. Enhanced connectivity within networks related to emotion regulation, cognitive control, and attention in responders suggests neural mechanisms supporting improved emotional and cognitive flexibility.

## Introduction

Affecting over 150 million people worldwide, major depressive disorder (MDD) is a second largest contributor to disability worldwide (Bromet et al., 2011; Organization, 2008). As the first-line of treatment, antidepressant drugs have become the cornerstone of treating MDD. Nevertheless, there is significant variability in treatment response and over a third of patients with MDD fail to respond to a given treatment (Trivedi et al., 2006) while approximately 40% develop treatment resistant depression where one does not remit after the prescription of two or more antidepressants (Souery et al., 2011). The primary barrier to improving treatment response is the lack of biomarkers that can predict an individual’s response to given treatment.

Functional magnetic resonance imaging (fMRI) is a powerful tool for exploring the pathophysiology of MDD, offering insights into brain circuit dysfunction and aiding the identification of biomarkers that could predict treatment response and track changes associated with successful interventions (Pilmeyer et al., 2022). Recent perspectives on MDD suggest that it is characterized by dysregulated neural networks rather than isolated disruptions in specific brain regions, including the default mode network (DMN), the salience network (SN), the cognitive control network (CCN), the affective network (AN), and parts of the limbic system (Janis Brakowski et al., 2017; Dutta et al., 2014; Long et al., 2020; Mulders et al., 2015).

Building on this network-based perspective, neuroimaging studies have increasingly focused on how these large-scale connectivity alterations relate to antidepressant treatment response (Chin Fatt et al., 2020; Demchenko et al., 2022; Qin et al., 2015; Saelens et al., 2024; Tozzi et al., 2024; Wang et al., 2024). Both task-based and resting-state functional MRI (rsfMRI) findings suggest that treatment efficacy is closely linked to functional interactions among these networks, particularly in regions implicated in emotional regulation and cognitive control (Li et al., 2022). Task-based fMRI studies indicate that early changes in emotional reactivity and cognitive control, particularly within fronto-limbic circuits, may predict later treatment response (Dunlop et al., 2017; Lai, 2021; Pizzagalli, 2011). In particular, responses to emotional stimuli in the amygdala and medial prefrontal cortex (mPFC) have been shown to normalize with successful treatment, suggesting a potential biomarker for antidepressant efficacy (Benedetta Vai et al., 2016). Pharmacological fMRI studies provide further evidence, demonstrating antidepressant modulation of neural activity even before clinical symptom improvement in key emotional processing regions, suggesting early neural changes preceding therapeutic effects (Chang et al., 2015; McCabe et al., 2011).

Altered between-network FC has been increasingly implicated in antidepressant treatment outcomes (Broeders et al., 2024; Fu et al., 2024; Sun et al., 2024). For example, alterations in dorsolateral prefrontal cortex (DLPFC) connectivity with the posterior DMN have also been associated with relapse following antidepressant discontinuation (Berwian et al., 2020). Similarly, in late-life depression, reduced RSFC within the CCN and DMN predicted lower remission rates and persistent symptoms such as apathy and dysexecutive behavior following escitalopram treatment (Alexopoulos et al., 2012). More recently, higher RSFC between the right fronto-parietal network (FPN) and posterior DMN, somatosensory network (SMN), and somatosensory association cortex has been linked to better treatment outcomes, emphasizing the role of network-level interactions in antidepressant efficacy (Kaiser et al., 2022; Martens et al., 2022).

Despite these advances, the clinical utility of RSFC findings remains limited by methodological variability, including differences in preprocessing pipelines, seed selection, and sample characteristics (Long et al., 2025; Prompiengchai & Dunlop, 2024; Tassone et al., 2024). Many studies have focused on either depressive symptomatology or treatment outcomes, often overlooking other clinically relevant dimensions such as cognitive impairment, which plays a crucial role in prognosis (Dichter et al., 2015; Hamilton et al., 2012). Additionally, while disruptions in fronto-limbic and DMN-CCN interactions are well-documented, their functional significance whether they primarily reflect depressive state, cognitive dysfunction, or treatment responsivity remains unclear. To address these gaps, this study employs a seed-voxel RSFC approach to examine the neural correlates of antidepressant response in MDD. By integrating RSFC analyses with validated clinical measures, we aim to clarify whether specific connectivity alterations serve as markers for treatment outcomes. Given prior findings linking RSFC patterns with antidepressant response, we hypothesize that antidepressant treatment will be preceded by distinct baseline functional connectivity (FC) patterns in networks involved in serotonin signaling and emotional processing, with greater FC at baseline observed in treatment responders compared to non-responders. With this approach, we aim to uncover neurobiological insights that could inform more personalized and effective antidepressant treatment strategies in MDD.

## Methods

### Participants and clinical assessments

A total of 86 patients with MDD (antidepressant responders, 39; non-responders, 47) and 93 HCs aging from 19-65 years and recruited between November 2016 and December 2021 from the outpatient psychiatric clinic of Korea University Anam Hospital in Seoul, Republic of Korea were included in the present study. All patients diagnosed with MDD and registered in this study received their diagnoses through the Structured Clinical Interview based on the Diagnostic and Statistical Manual of Mental Disorders, Fifth Edition(First et al., 2016), conducted by two experienced board-certified psychiatrists (K.-M. Han and B.-J. Ham). The following exclusion criteria were adopted to the MDD group: (i) comorbidity of any other major psychiatric disorders (including personality and substance use disorders), (ii) MDD with psychotic features, (iii) acute suicidal or homicidal patients who require immediate inpatient treatment, (iv) history of severe or unstable medical conditions that can affect the immune system, (v) underlying primary neurological illness (i.e., Parkinson’s disease, cerebrovascular disease, or epilepsy), (vi) abnormal results on recent physical examination or laboratory tests, (vii) pregnancy or currently nursing, and (viii) any factors that would hinder MRI. HC group were recruited from the community through advertisements and were assessed by the two psychiatrists with the identical exclusion criteria as those used for the MDD group. Additional confirmation from the two board-certified psychiatrists was required for participants to be included, ensuring that they had no ongoing or past history of mental disorders.

Sociodemographic and clinical information, including age, sex, years of education were collected from both groups. The severity of depressive symptoms among all participants was evaluated using the 17-item Hamilton Depression Rating Scale (HDRS) by psychiatrists at the day of the MRI scan(Hamilton, 1959, 1960). Antidepressant response was defined as ≥50% improvement after 8 weeks of pharmacological treatment from the total HDRS score at baseline. While the majority of participants received monotherapy, medication switches were allowed when clinically necessary. The use of psychopharmacological treatments at the time of the MRI scan was assessed to account for the possible influence of them. More details of the information are shown in Table 1.

**Table 1.**
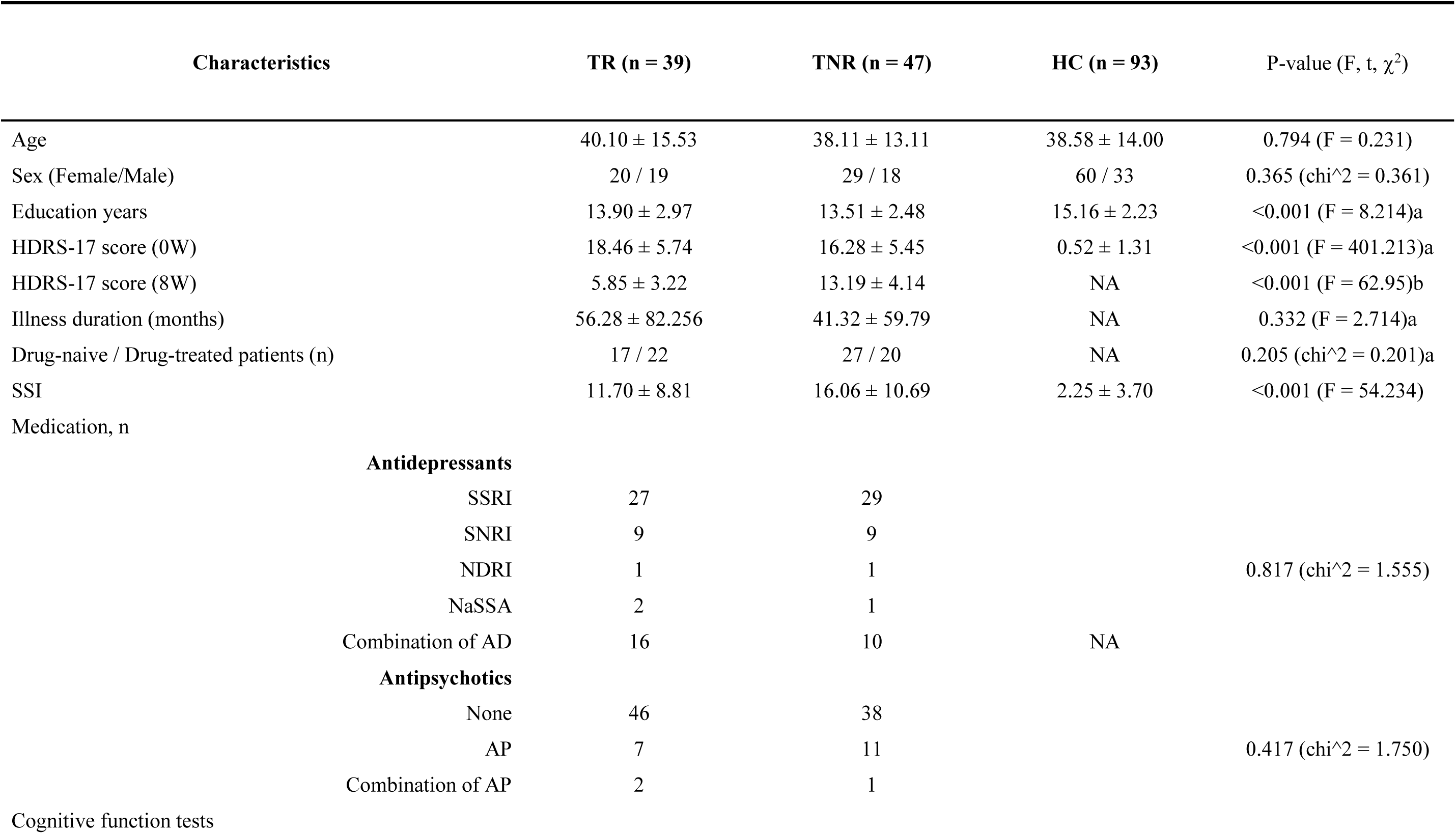
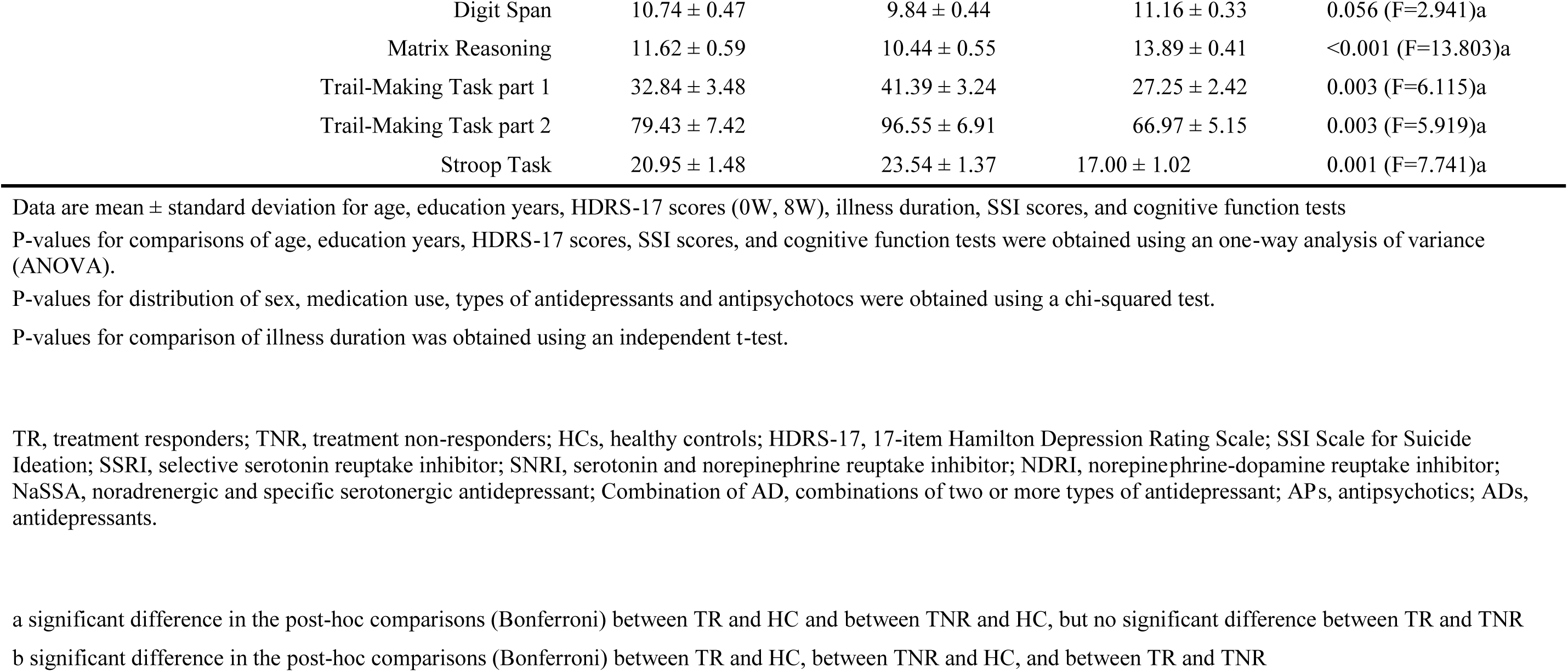
Demographic and clinical characteristics of patients with major depressive disorder and healthy controls.

The study was conducted in accordance with the ethical principles of the Declaration of Helsinki approved by the Institutional Review Board of Korea University Anam Hospital (IRB No. 2019AN0174). All participants provided written voluntary consent and signed informed documentation, recognizing their right to withdraw from the study at any time.

### Cognitive test battery

To evaluate the cognitive abilities of participants, we employed a combination of tasks, including the digit span, matrix reasoning, trail-making, and Stroop tests, which assessed various domains such as attention, nonverbal reasoning, mental flexibility, and executive functions. All assessments were conducted in Korean-speaking environments.

The digit span and matrix reasoning tasks were derived from the Wechsler Adult Intelligence Scale (WAIS) (WECHSLER, 1958). The digit span test, used to measure verbal IQ and working memory, involved three components: repeating numbers as presented, recalling numbers in reverse order, and reorganizing them in ascending order. For nonverbal IQ, the matrix reasoning task assessed perceptual reasoning by asking participants to identify the correct completion for a series of incomplete visual patterns. To control for age-related variability, the total scores for the digit span and matrix reasoning tasks were adjusted based on participants’ ages.

The trail-making test, originally part of the Army Individual Test of General Ability(Reitan, 1955), was later adapted for evaluating cognitive functions in individuals with brain damage and remains widely used. Participants are instructed to connect numbers from 1 to 25, scattered randomly on paper, as quickly as possible without stopping. Then, they are asked to do the same for alternating numbers and letters in ascending order (e.g., 1-A-2-B-3-C-4-D…). The Stroop test (Stroop, 1935) followed the standard procedure, with a maximum possible score of 26 consisting of three components – naming colors, color of words unrelated to colors, and color in which a conflicting color word is written. Performance on both the trail-making and stroop tests was evaluated based on reaction time and the number of errors.

### fMRI data acquisition

All the participants underwent the MRI scan at the day of the blood sampling and clinical assessments. Functional and T1-weighted images of the participants were obtained using a 3.0-Tesla TrioTM whole-body imaging system (Siemens Healthcare GmbH, Erlangen, Germany) at the Korea University MRI Center. T1-weighted images were acquired using the MPRAGE sequence (TR = 1900 ms, TE = 2.52 ms, flip angle = 9°, matrix = 256 × 256, resolution = 1 × 1 × 1 mm^3^) covering the whole head. Functional images were obtained using a gradient-echo echo planar imaging (EPI) sequence (TR = 2000 ms, TE = 30 ms, flip angle = 90°, number of slices = 75, matrix = 80 × 80, resolution = 2 × 2 × 2 mm^3^, multiband acceleration factor = 3, 155 volumes). During the scan, participants were instructed to keep their eyes closed, remain still, and not to fall asleep. All the images were checked by visual inspection for gross motion artifacts and potential artifacts due to metallic foreign bodies, and there were no images to be discarded after visual inspection.

### fMRI data preprocessing

CONN functional connectivity toolbox (Version 22a) (Nieto-Castanon & Whitfield-Gabrieli, 2012) and SPM12 (http://www.fil.oin.ucl.ac.uk/spm) were used to preprocess the resting-state fMRI (rsfMRI) of the participants. Flexible pipeline implemented in the CONN Toolbox was used to preprocess the functional and anatomical data according to the methods described in previous studies (Nieto-Castanon, 2020). In summary, the pipeline included the following preprocessing steps: slice timing correction, outlier identification, direct segmentation and MNI-space normalization, realignment with correction of susceptibility distortion interactions, and smoothing. Realigning of the functional data using the SPM realign & unwarp procedure (Andersson et al., 2001), and then the coregistration of all scans to the first scan of the first session using a least squares method and a six-parameter (rigid body) transformation was performed (Friston et al., 1995). The preprocessing includes detection of potential outlier scans using the Artifact Detection Tools scrubbing procedure as acquisitions with framewise displacement over 0.9 mm or global BOLD signal changes above 5 standard deviations (Nieto-Castanon & Whitfield-Gabrieli). After a direct normalization procedure, functional and anatomical data underwent following processes: normalization into standard MNI space, segmentation into grey matter, white matter, and CSF tissue classes, and resampling to 2 mm isotropic voxels using the SPM algorithm. Finally, smoothing of the functional data was performed using spatial convolution and a Gaussian kernel with an 5 mm full width half maximum (FWHM). In addition to these processes, a standard denoising pipeline was applied to denoise the functional data with the component-based noise correction (CompCor) (Nieto-Castanon, 2020), and the bandpass frequency filter between 0.008 Hz and 0.09 Hz was applied to BOLD timeseries to minimize the noise generated by scanner drift and physiological effects (Hallquist et al., 2013).

### Functional connectivity analysis

For the comparison of the RSFC between the three groups, the seed-to-voxel functional connectivity (FC) analysis and ROI-to-ROI analysis were performed using CONN functional connectivity toolbox (Version 22a) (Nieto-Castanon and Whitfield-Gabrieli, 2012). For the analyses, we used the brain regions consists of the hubs of the default mode network (DMN), salience network, cognitive control network (CCN), and limbic network based on previous conceptual frameworks of neural networks involved in the pathophysiology of depression by Williams (2016) and by Phillips (2015) as the seeds. Notably, these seed regions also overlap with cortical areas showing high serotonergic receptor binding, supporting the biological plausibility of our hypothesis regarding serotonin-modulated network dynamics (Hansen et al., 2022). The following ROIs were selected as seed regions from suggestion of previous studies on RSFC studies and MDD as follows: left subgenual anterior cingulate cortex (sgACC) (-4, 26, -10) (Hamani et al., 2011; Pizzagalli, 2011), anterior cingulate cortex (ACC) (0, 48, -4) (Lui et al., 2011; van der Wijk et al., 2021), left ventral anterior cingulate cortex (vACC) (-3, 33, 11) (Zhu et al., 2012), posterior cingulate cortex (PCC) (±11, -52, 37/35) (Goldstein-Piekarski et al., 2018), left precuneus (-6, -70, 62) (Alexopoulos et al., 2012), ventromedial prefrontal cortex (VMPFC) (±4, 36, -16) (Alexopoulos et al., 2012; Li et al., 2013), medial prefrontal cortex (mPFC) (±2, 50, -6) (Goldstein-Piekarski et al., 2018; Zhu et al., 2012), rostral/pregenual anterior cingulate cortex (rpACC) (0, 39, 10) (Amit Anand et al., 2007; Critchley, 2004; Damasio, 1997), dorsal anterior cingulate cortex (dACC) (±4, 30, 22) (Alexopoulos et al., 2012), dorsolateral prefrontal cortex (DLPFC) (±52, 28, 28) (van der Wijk et al., 2021), thalamus (±11, -16, 5) (Amit Anand et al., 2007; Lui et al., 2011), amygdala (±17, -5, -12) (Amit Anand et al., 2007; Kozel et al., 2011), right orbitofrontal cortex (12, 36, -20) (Kozel et al., 2011), and insula (±40, 20, 0) (Lui et al., 2011; van der Wijk et al., 2021). The ROIs were defined as spheres of 10 mm in radius and were centered at MNI coordinates according to the Harvard-Oxford Subcortical Structural Probability Atlas in Functional Magnetic Resonance Imaging of the Brain Software Library.

### Statistical analysis

For the investigation of the RSFC differences between antidepressant responders and non-responders, we primarily performed comparison of the FC between the three groups (antidepressant responders, non-responders, HC) using the seed-to-voxel and ROI-to-ROI FC analysis. We used the mean time series of each seed as a predictor in a multiple regression general linear model (GLM) at each voxel, and then the Pearson’s correlation analyses were performed including all other voxels in the whole brain. Using the Fisher’s r-to-z transformation, Pearson’s correlation coefficients were converted to z-scores, which were then used in the group level analyses. To compare the FC between the groups, GLM was adopted to analyze the group differences in FC maps between the three groups including age, sex, and education years as nuisance covariates in all the analyses. A GLM was performed with the independent t-test to compare the mean z-scores of two groups. Statistically significant seed to cluster FC was determined as follows: voxel-wise uncorrected P < 0.001 to define the voxel size, followed by a cluster-wise correction was applied at a threshold of false discovery rate (FDR)-corrected P < 0.05 within each seed-to-voxel analysis to determine significant clusters (minimum cluster size: 20). Then, the mean z-scores (i.e., a measure of FC) of statistically significant seed to cluster FC were extracted using the CONN toolbox. As the second part of the analyses, for the correlation analyses between FC, cognitive scores, and illness severity were performed in the MDD (responder and non-responder) and HC group, respectively. We conducted Pearson’s correlation analyses between mean z-scores of the significant seed-to-cluster FC and cognitive scores including age, sex, education years, and illness duration (only for the MDD group) as nuisance covariates with the statistical threshold of P < 0.05. False Discovery Rate (FDR) correction was again applied separately to adjust for the multiple comparison effect.

For the comparison of the sociodemographic and clinical characteristics between the three groups, independent t test was used to analyze age, education years, psychiatric scales, and total intracranial cavity volume (cm^3^), and the chi-square test to analyze the differences in sex. Above-mentioned statistical analyses and the correlation analyses were performed using IBM SPSS Statistics for Windows (version 25.0; IBM Corp., Armonk, NY, USA).

## Results

### Demographic and clinical characteristics of the participants

The demographic and clinical characteristics of the participants are summarized in Table 1. There were no significant differences between the MDD and HC groups in terms of age, sex, years of education, and TICV. Nevertheless, the MDD group exhibited significantly higher HDRS score than the HC group (P < 0.001). Additionally, while no significant difference was observed in the baseline HDRS score between treatment responders and non-responders, the HDRS score after 8 weeks of treatment in responders was significantly higher than that of non-responders (p<0.001). Among the 86 patients with MDD, 44 (51.2%) were drug-naïve and 42 (48.8%) were taking psychotropic medications during blood sample acquisition and MRI scanning, and there was no significant difference in the medication status between treatment responders and non-responders.

In cognitive function assessments, antidepressant non-responders demonstrated the lowest overall scores, while the HCs achieved the highest (Table 1). Compared to HCs, patients with MDD exhibited significantly lower performance across all cognitive tasks, except for the digit span task (P=0.056). Post-hoc Bonferroni analysis indicated that both MDD subgroups performed significantly worse on all cognitive tasks compared to HCs.

### Differences in resting-state functional connectivity across Major Depressive Disorder and between antidepressant responders and non-responders

From the seed-based analysis, MDD patients showed enhanced connectivity in the ACC with large clusters in regions such as the PCC, DLPFC, thalamus, and occipital cortex. In contrast, the PCC revealed reduced functional connectivity in MDD patients compared to HC, particularly in the opercular cortex, precentral gyrus, and inferior frontal gyrus, with altered connectivity in the PCC, angular gyri, and occipital cortex. Additionally, both the amygdala and thalamus demonstrated significant differences, with altered connectivity patterns in regions including the prefrontal cortex, occipital cortex, and parietal lobe (Supplementary Table 1).

Additionally, compared with antidepressant non-responders (TNR), antidepressant responders (TR) exhibited significantly higher FCs between the following seeds and clusters: the left DLPFC and left lingual gyrus; left PCC and cluster comprising the precuneus and the right cerebellum; the left VMPFC and lateral occipital cortices; left amygdala and cluster comprising right lateral occipital cortex and left supramarginal gyrus; right amygdala and cluster comprising precuneus and left lateral occipital cortex; left dACC and precuneus; left sgACC and left lateral occipital cortex; left thalamus and cluster comprising right middle frontal/temporal gyrus, postcentral gyrus, left superior frontal gyrus, right lateral occipital cortex, and left temporal pole; right thalamus and cluster comprising precentral gyrus, left superior frontal gyrus, right middle frontal gyrus, and right postcentral gyrus (Table 2, Fig 1). The responder group further showed significantly higher FCs between the left PCC and left sgACC compared to those in the non-responder group (FDR-corrected P-value = 0.009), with a medium effect size (Cohen’s d = 0.51) in the ROI-to-ROI analysis (Table 3, Fig 2).

**Fig 1.**
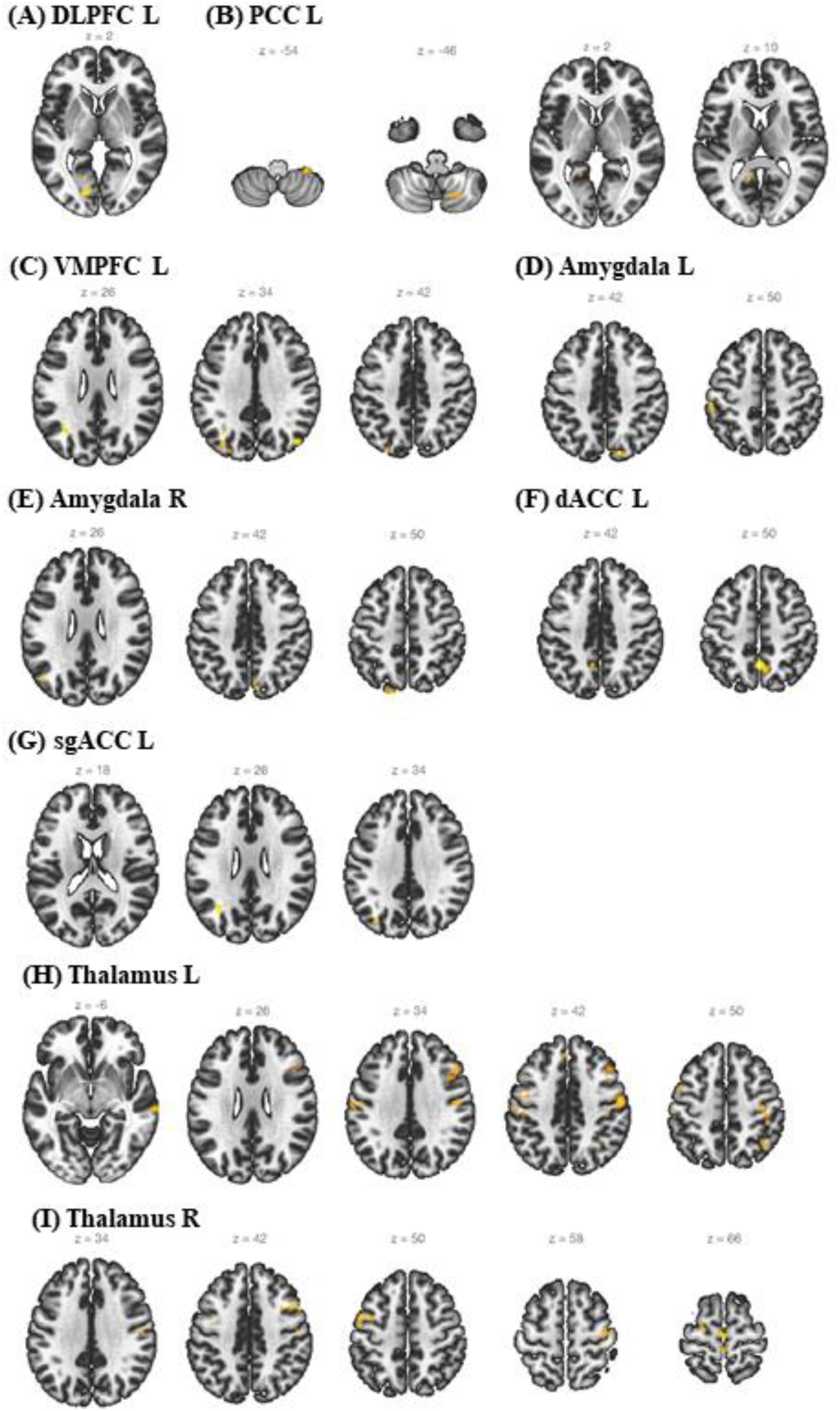
Group differences in functional connectivity. Axial views of the different brain regions show significantly higher functional connectivity (FC) in treatment responders (TR) compared to non-responders (TNR) with the following seeds: (A) the left dorsolateral prefrontal cortex (DLPFC L), (B) the left posterior cingulate cortex (PCC L), (C) the left ventromedial prefrontal cortex (VMPFC L), (D) the left amygdala (Amygdala L), (E) the right amygdala (Amygdala R), (F) the left dorsal anterior cingulate cortex (dACC L), (G) the left subgenual anterior cingulate cortex (sgACC L), (H) the left thalamus (Thalamus L), (I) the right thalamus (Thalamus R).

**Fig 2.**
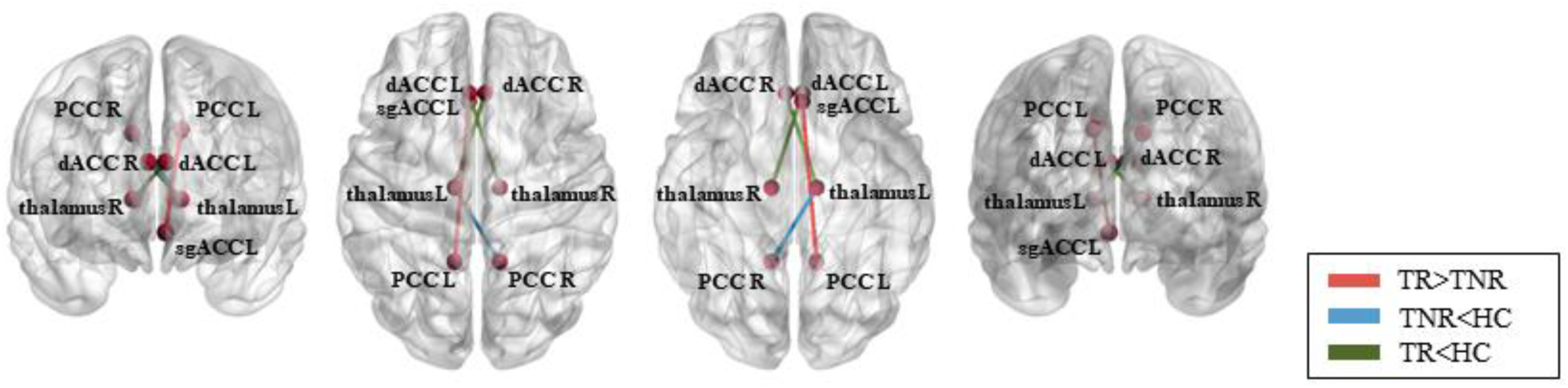
Group differences in functional connectivity – ROI-to-ROI. The different brain regions showing significant differences in functional connectivity (FC) between treatment responders (TR), non-responders (TNR), and healthy controls (HC).

**Table 2.**
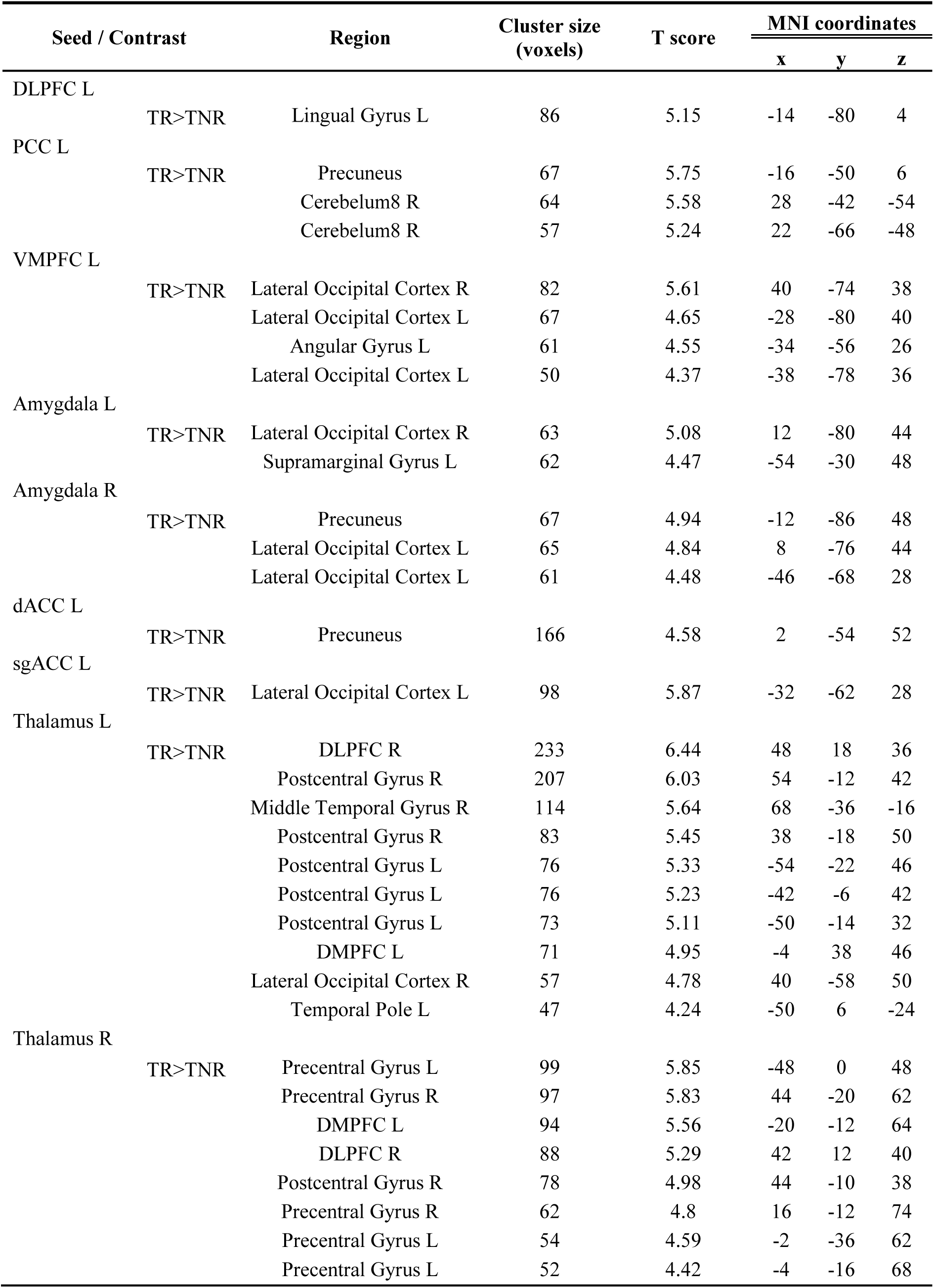
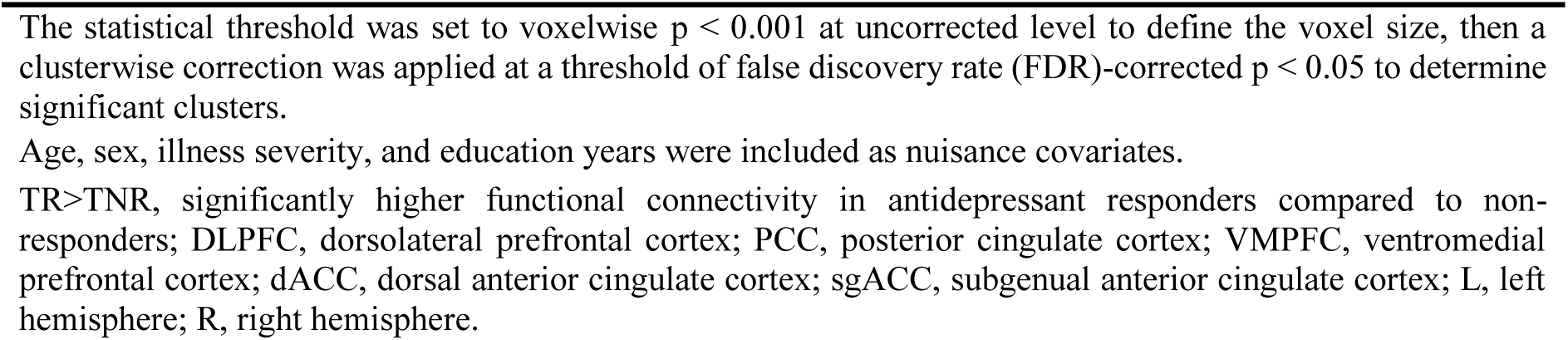
Seed-to-Voxel analysis between patients with treatment responders (TR) and non-responders (TNR).

**Table 3.** ROI-to-ROI analysis between patients with major depressive disorder (MDD) and healthy controls (HC)

| **Brain regions** | **Contrast** | **FDR-corrected P-value** |
| --- | --- | --- |
| PCC L - sgACC L | TR>TNR | 0.009 |
| PCC R - thalamus L | TNR<HC | 0.043 |
| dACC L - thalamus R | TR<HC | 0.041 |
| dACC R - thalamus L | TR<HC | 0.038 |

Age, sex, illness severity, and education years were included as nuisance covariates.
TR>TNR, significantly higher functional connectivity in antidepressant responders compared to non-responders; TNR<HC, significantly lower functional connectivity in antidepressant non-responders compared to healthy controls; TR<HC, significantly lower functional connectivity in antidepressant responders compared to healthy controls; PCC, posterior cingulate cortex; dACC, dorsal anterior cingulate cortex; sgACC, subgenual anterior cingulate cortex; L, left hemisphere; R, right hemisphere; pFDR, FDR-corrected P-value.

To further contextualize these findings and address deviations from normative connectivity patterns, we compared TR and TNR to HC. Compared to HC, TRs exhibited significantly higher functional connectivity between the VMPFC and dorsolateral prefrontal cortex, amygdala and lateral occipital cortex, PCC and cerebellum, and thalamus and postcentral gyrus (Supplementary Table 2). In contrast, TNRs showed significantly lower connectivity than HC across widespread regions, including the ACC, PCC, VMPFC, and mPFC, with the angular gyrus, hippocampus, temporal poles, and occipitoparietal cortices (Supplementary Table 3).

### Correlations between cognitive scores and functional connectivity in the responders and non-responders

Regarding the correlation analyses, we extracted the mean z-scores of the statistically significant seeds-to-cluster FC based on the results of the FC comparison between the two groups.

In the responder group, we observed a significant negative correlation of the FC between the left amygdala and left supramarginal cortex, digit span task (r = -0.350, P = 0.044), and matrix reasoning task (r = -0.451, P = 0.022). Similarly, trail-making task part 2 was also negatively correlated with FC between the left VMPFC and left lateral occipital cortex (r = -0.364, P = 0.044); right thalamus and right precentral gyrus (r = -0.369, P = 0.044). We further observed a significant positive correlation of the FC between the left thalamus and left postcentral gyrus (r = 0.373, P = 0.044) and trail-making task part 1. The FC between the right thalamus and right middle frontal gyrus was positively correlated with the Stroop task (r = 0.429, P = 0.025) and trail-making task part 2 (r = 0.375, P = 0.044). The Stroop task was also positively correlated with the FC between the right thalamus and right precentral gyrus (r = 0.355, P = 0.044) (Table 4, Fig 3).

**Fig 3.**
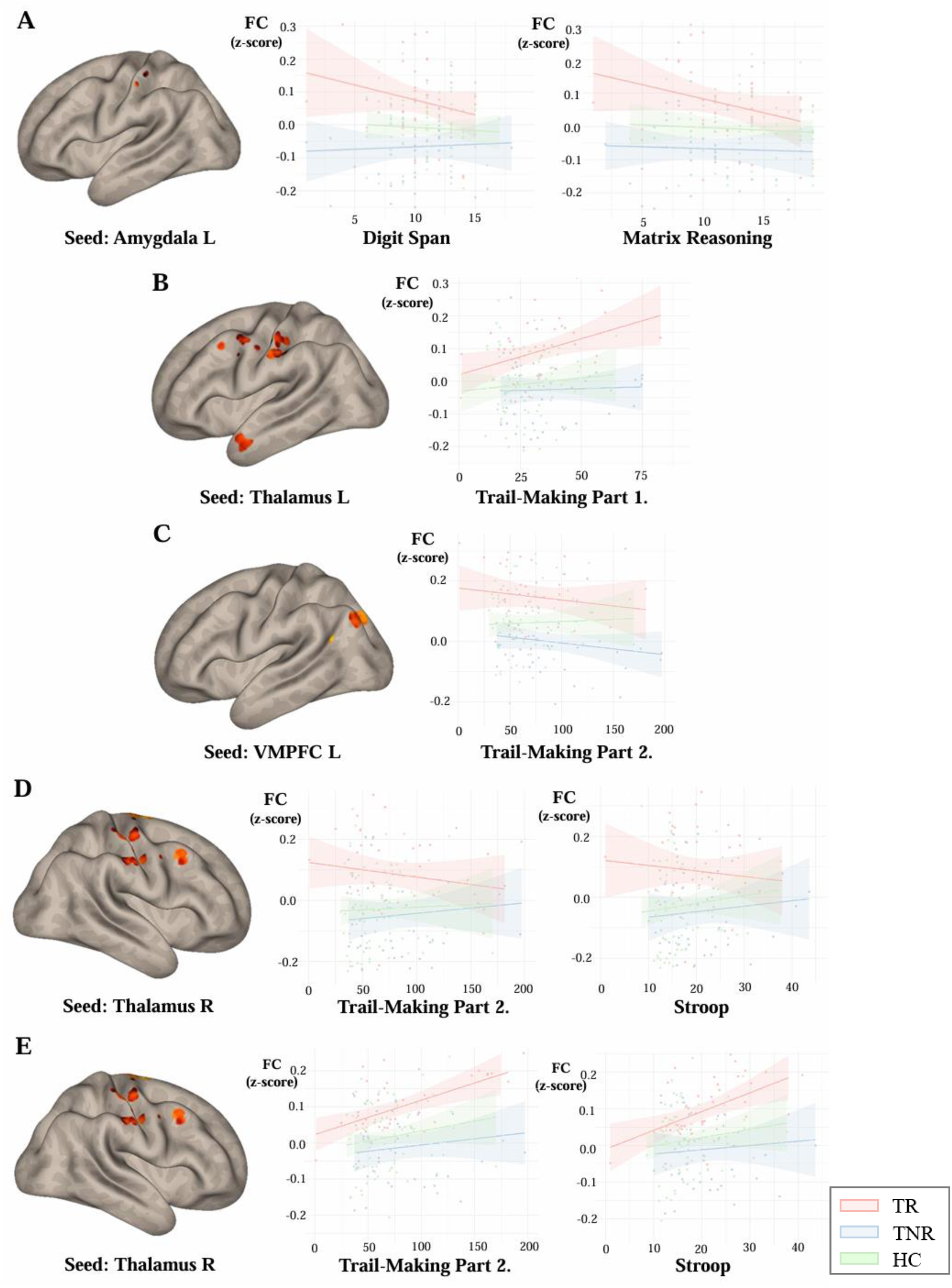
Correlations between cognitive function and functional connectivity (FC) among patients with major depressive disorder (MDD) and healthy controls (HC). Panel A: FC between the left amygdala and clusters including the left supramarginal cortex, along with scatter plots illustrating its correlation with digit span and matrix reasoning tasks, respectively; Panel B: FC between the left thalamus and clusters including the left postcentral gyrus, with a scatter plot depicting its correlation with trail-making task part 1; Panel C: FC between the left ventromedial prefrontal cortex (VMPFC) and clusters including the left lateral occipital cortex, along with a scatter plot showing its correlation with trail-making task part 2; Panel D: FC between the right thalamus and clusters including the right precentral gyrus, with scatter plots illustrating its correlation with trail-making task part 2 and stroop task, respectively; Panel E: FC between the right thalamus and clusters including the right middlefrontal gyrus, with scatter plots showing its correlation with trail-making task part 2 and stroop task, respectively; TR, treatment responder; TNR, treatment non-responder; HC, healthy control; FC, functional connectivity; VMPFC, ventromedial prefrontal cortex; L, left hemisphere; R, right hemisphere.

**Table 4.** Correlation analysis between functional connectivity and cognitive function in treatment responders.

| **Cognitive task** | **Region** | **r** | **p FDR-corrected** |
| --- | --- | --- | --- |
| Digit Span | Amygdala L - Supramarginal Cortex L | -0.350 | 0.044 |
| Matrix Reasoning | Amygdala L - Supramarginal Cortex L | -0.451 | 0.022 |
| TMT_1 | Thalamus L - postcentral gyrus L | 0.373 | 0.044 |
| TMT_2 | VMPFC L - Lateral Occipital Cortex L | -0.364 | 0.044 |
|  | Thalamus R - precentral gyrus R | -0.369 | 0.044 |
|  | Thalamus R - middlefrontal gyrus R | 0.375 | 0.044 |
| Stroop Task | Thalamus R - middlefrontal gyrus R | 0.429 | 0.025 |
|  | Thalamus R - precentral gyrus R | 0.355 | 0.044 |

Pearson's partial correlation analyses between connectivity strength (mean z-value) and cognitive tasks within the TR group.
Age, sex, education years, illness duration were included as nuisance covariates.
VMPFC, ventromedial prefrontal cortex; TMT\_1, trail-making task part 1; TMT\_2, trail-making task part 2; L, left hemisphere; R, right hemisphere; pFDR, FDR-corrected P-value.

In the non-responder group, the FC between the left sgACC and left lateral occipital cortex was negatively correlated with digit span task (r = -0.479, P = 0.006), while being positively correlated with matrix reasoning task (r = 0.424, P = 0.014), trail-making task part 1 (r = 0.486, P = 0.003) and 2 (r = 0.581, P = 0.001), and the Stroop task (r = 0.532, P = 0.002). Additionally, we observed significant negative correlation between matrix reasoning task and FC between the following seeds and clusters: left amygdala and right lateral occipital cortex (r = -0.324, P = 0.044); right amygdala and precuneus (r = -0.336, P = 0.044). The FC between the left VMPFC and left lateral occipital cortex was negatively correlated with matrix reasoning task (r = -0.325, P = 0.044) while being positively correlated with trail-making task part 1 (r = 0.313, P = 0.050), 2 (r = 0.388, P = 0.025), and the Stroop task (r = 0.352, P = 0.039). Similarly, trail-making task part 2 was also positively related with FC between left VMPFC and right lateral occipital cortex (r = 0.482, P = 0.003) (Supplementary Table 4).

We have also identified moderate correlations between FCs and the HDRS scores in the following seeds and clusters: right amygdala and precuneus (r = 0.330, P = 0.050); right thalamus with bilateral precentral cortices (L: r = 0.335, P = 0.050; R: r = 0.425, P = 0.050), and left superiorfrontal cortex (r = 0.345, P = 0.050) in the responder group (Table 5, Fig 4). All P-values presented here reflect FDR-corrected results.

**Fig 4.**
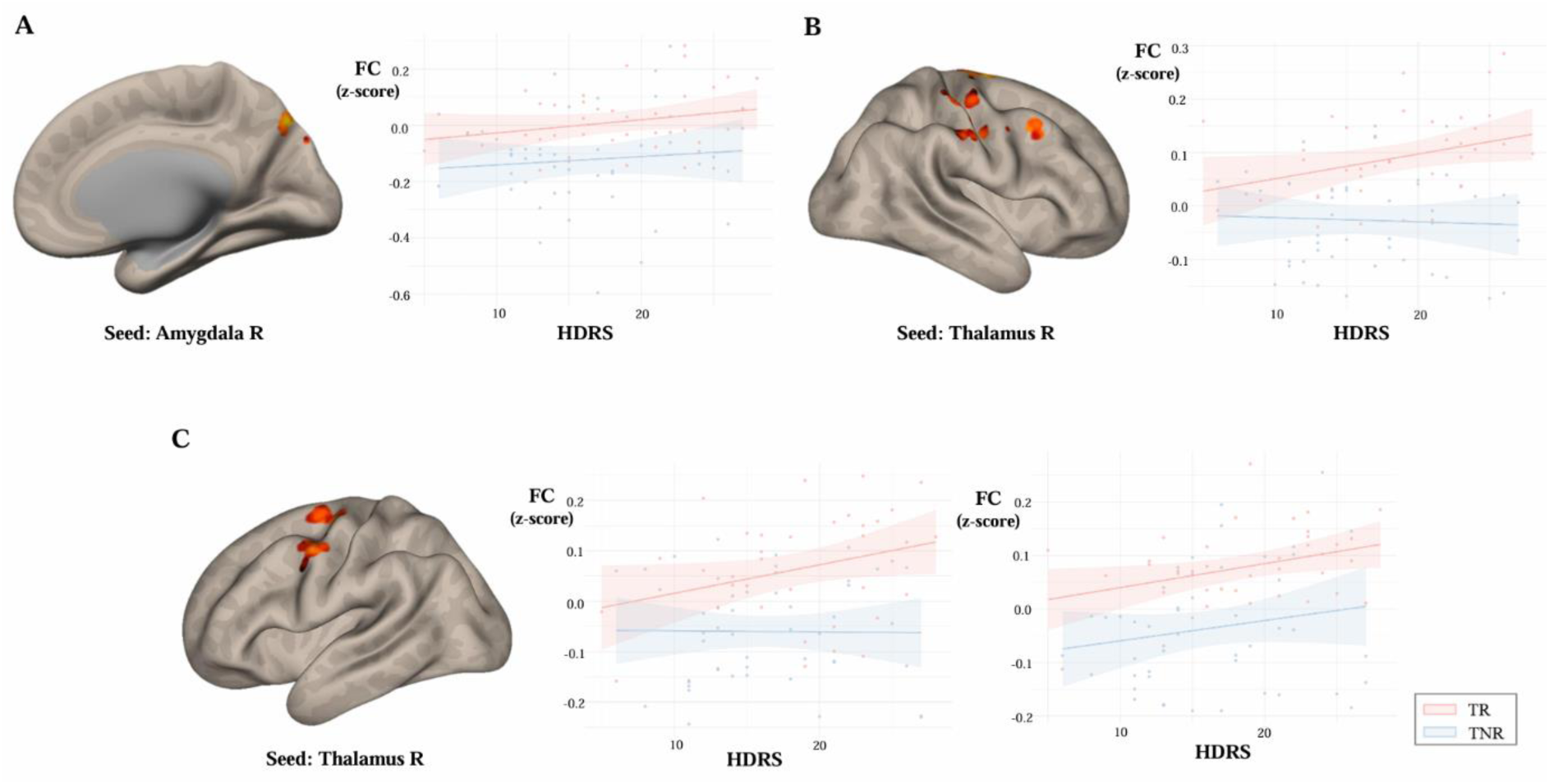
Correlations between illness severity and functional connectivity (FC) among patients with major depressive disorder (MDD). Panel A: FC between the right amygdala and clusters including precuneus, along with a scatter plot illustrating its correlation with the 17-item Hamilton Depression Rating Scale (HDRS); Panel B: FC between the right thalamus and clusters including the right precentral cortex, with a scatter plot depicting its correlation with HDRS; Panel C: FC between the right thalamus and clusters including the left precentral cortex and left superior frontal cortex, as well as scatter plots showing their respective correlations with HDRS (left precentral cortex and left superior fontal cortex, respectively); TR, treatment responder; TNR, treatment non-responder; HDRS, 17-item Hamilton Depression Rating Scale; FC, functional connectivity; L, left hemisphere; R, right hemisphere.

**Table 5.** Correlation analysis between functional connectivity and illness severity.

| **group** | **Region** | **R** | **p FDR-corrected** |
| --- | --- | --- | --- |
| TR | Amygdala R - precuneus | 0.33 | 0.050 |
|  | Thalamus R - precentral cortex L | 0.335 | 0.050 |
|  | Thalamus R - superiorfrontal L | 0.345 | 0.050 |
|  | Thalamus R - precentral cortex R | 0.425 | 0.050 |
| TNR | sgACC - lateral occipital cortex L | 0.354 | 0.050 |
| HC | Amygdala R - lateral occipital cortex L | -0.233 | 0.050 |
|  | Thalamus L - Postcentral gyrus L | -0.27 | 0.050 |

Pearson's partial correlation analyses between connectivity strength (mean z-value) and illness severity within each groups.
Age, sex, education years, illness duration were included as nuisance covariates.
TR, treatment responders; TNR, treatment non-responders; HC, healthy controls; sgACC, subgenual anterior cingulate cortex; L, left hemisphere; R, right hemisphere; pFDR, FDR-corrected P-value.

## Discussion

Identifying reliable biomarkers for antidepressant response is critical for improving treatment outcomes in MDD, given the heterogeneous nature of the condition and the variability in treatment efficacy. The current study, therefore, aimed to investigate not only the underlying RSFC differences in MDD but also their associations with critical aspects of the disorder that influence prognosis and treatment outcomes, including depressive state and cognitive impairment. Our seed-based analysis revealed that antidepressant responders exhibited significantly higher FCs between specific brain regions compared to non-responders, particularly involving the prefrontal cortex, thalamus, amygdala, and PCC. Furthermore, several interregional FCs were significantly correlated with depressive severity and cognitive task performance, suggesting that RSFC patterns may reflect significant correlations between antidepressant response, depressive state, and cognitive functions. Additional comparisons with HCs revealed that TRs showed relatively increased FC in similar regions, while TNRs showed more widespread reductions across frontoparietal and limbic areas, consistent with the pattern of disrupted network integration observed in the TR and TNR comparisons.

RSFC findings in MDD have been highly heterogeneous, with studies reporting both hypo- and hyperconnectivity depending on the networks examined and methodological differences (Kaiser et al., 2015; Mulders et al., 2015; Veer et al., 2010). In line with this complexity, our study revealed a mixed pattern of alterations, with significant hyperconnectivity in the ACC with large clusters in the PCC, DLPFC, thalamus, and occipital cortex, alongside reduced functional connectivity of the PCC with the opercular cortex, precentral gyrus, and inferior frontal gyrus. Additionally, the amygdala and thalamus exhibited altered connectivity with prefrontal, occipital, and parietal regions. Increased ACC connectivity with cognitive control and default mode regions aligns with models proposing that MDD involves excessive self-referential processing, sustained negative affect regulation, and disrupted network balance(Kaiser et al., 2015; Mulders et al., 2015). The hyperconnectivity of the ACC, PCC, and DLPFC may therefore, be an indication of persistent recruitment of cognitive control regions in an effort to regulate mood, albeit ineffectively(Hamilton et al., 2011; Pizzagalli, 2011). Conversely, reduced PCC activation reflects dysfunction within the DMN, consistent with prior findings linking PCC hypoactivity to maladaptive self-focus and rumination(Bartova et al., 2015; Nejad et al., 2013). Moreover, heightened connectivity in the amygdala and thalamus suggests dysregulated emotional and sensory processing, which may contribute to the emotional dysregulation characteristic of MDD. These findings also align with growing evidence implicating the thalamus in the pathophysiology of depression and its potential role in treatment response (J. Brakowski et al., 2017; Yamamura et al., 2016; Yang et al., 2023). Altered thalamic activity has been consistently reported in MDD, with evidence suggesting that disruptions in thalamocortical connectivity and excessive bottom-up signaling from the thalamus, alongside weakened top-down control, may contribute to both affective dysregulation and cognitive dysfunction (Li et al., 2013; Zhou et al., 2020).

In contrast to these baseline MDD findings, when comparing antidepressant responders to non-responders, responders exhibited significantly higher FC between regions involved in emotion regulation, attention, and cognitive control. These included connections between the DLPFC and visual processing regions (e.g., lingual gyrus), the PCC and areas within the DMN (e.g., precuneus and cerebellum), as well as the VMPFC and the occipitoparietal regions. Enhanced FC was also observed between the amygdala, cingulate cortices (dACC and sgACC), and regions associated with the SN and sensorimotor processing (e.g., occipital cortex, supramarginal gyrus, and thalamus). These findings align with prior research demonstrating that antidepressant response is closely associated with connectivity patterns within networks governing emotion regulation, cognitive control, and attention (J. Brakowski et al., 2017; Chin Fatt et al., 2020; DeMaster et al., 2022; Qin et al., 2015; Q. Wang et al., 2022). The therapeutic effects of antidepressants have been linked with their ability to modulate neuronal excitability, increase synaptic translation and release of BDNF, activate of signaling pathways, and promote synaptogenesis within the prefrontal cortices(Björkholm & Monteggia, 2016; Duman & Li, 2012; Wang et al., 2008). Such neurobiological changes may enhance FC across networks, including the cerebellum, DMN, affective networks, and SMN, which are increasingly recognized as playing pivotal roles in MDD (Qin et al., 2015; Rantamäki & Yalcin, 2016). In this context, the observed enhanced FC between the amygdala and regions like the cingulate cortices, sensorimotor areas, and components of the SN in responders underscores the integrative role of these networks in mediating the antidepressant response. Rather than reflecting changes induced by treatment, these patterns may represent pre-existing neurobiological features that support treatment efficacy. This further suggests that the interplay between network-level connectivity and biological readiness may underlie improvements in both emotional regulation and cognitive control, facilitating recovery in MDD (Zhou et al., 2022).

The Establishing Moderators and Biosignatures of Antidepressant Response in Clinical Care (EMBARC) study further supports the role of intrinsic FC patterns in predicting antidepressant response (Chin Fatt et al., 2020). In this study, baseline structural and rsfMRI were analyzed in 279 participants randomized to 8 weeks of sertraline or placebo treatment. Higher within-network connectivity in the DMN and greater between-network connectivity of the DMN and executive control networks (ECN) predicted better outcomes specifically for sertraline. Additionally, in a magnetoencephalography study of 66 medication-free patients with MDD, beta power in the SN significantly differed between therapeutically effective and ineffective groups and FC between the DMN, SN, and ECN correlated with treatment outcomes, with receiver operating characteristic (ROC) analysis yielding strong predictive accuracy (Q. Wang et al., 2022). These findings converge with prior research emphasizing the predictive value of DMN connectivity (Baeken et al., 2017; Dunlop et al., 2017; Kozel et al., 2011). For instance, stronger baseline PCC-mPFC connectivity has been reported to predict response to both pharmacotherapy and electroconvulsive therapy (ECT), with classification accuracies exceeding 80% in initial studies (Salomons et al., 2014), although such high predictive accuracy has not been consistently replicated in subsequent research. More recently, a cross-trial machine learning study using data from both the EMBARC and CAN-BIND cohorts demonstrated that pretreatment functional connectivity features, particularly in the dorsal ACC, modestly predicted antidepressant response, highlighting the importance of validating biomarkers across heterogeneous clinical populations (Zhukovsky et al., 2025).

In parallel, chronic stress-induced models of depression in rodents have consistently demonstrated the relationship between antidepressant treatments and neuroplasticity in key limbic regions implicated in both cognitive function and mood regulation (Elizalde et al., 2010; Haenisch et al., 2009; Hare et al., 2017; Yu et al., 2016). Several of these studies have revealed that chronic administration of antidepressants decreases excitatory neurotransmission and restores inhibitory control within the amygdala, contributing to improved emotional regulation (Pehrson et al., 2022; J. Zhou et al., 2014; W. Zhou et al., 2014). These effects align with our findings highlighting enhanced FC between the amygdala and prefrontal regions in antidepressant responders, potentially reflecting improved top-down regulation of limbic hyperactivity (Huang et al., 2014; Ma, 2015; B. Vai et al., 2016). These studies also provide a biological framework for understanding how antidepressant-induced modulation of the limbic system may underpin the observed FC changes in MDD.

Together, these findings underscore the interplay between intrinsic FC patterns across networks governing emotion, cognition, and salience detection. Furthermore, they suggest that antidepressant responders exhibit better integration between the DMN, SN, and frontoparietal control network (FPCN), which may support improved emotional and cognitive flexibility essential for treatment success. However, it remains an open question whether these connectivity patterns represent inherent traits that predispose individuals to respond favorably to treatment or are themselves modulated by the pharmacological effects of antidepressants. This accentuates the importance of further research to disentangle these relationships and better understand the interplay between intrinsic brain activity and treatment response.

Notably, in the responder group, FCs between regions such as the thalamus, amygdala, and lateral occipital cortex were associated with cognitive tasks, including the digit span, matrix reasoning, trail-making, and Stroop tasks, with both positive and negative correlations observed depending on the regions involved. In contrast, the non-responder group showed distinct FC – cognitive function relationships, particularly with connections involving the sgACC, VMPFC, and thalamus. Studies have consistently highlighted the close relationship between emotional regulation and cognitive function in patients with mood disorders, emphasizing how deficits in cognitive control disrupt the interplay, thereby creating a cycle of maladaptive strategies and cognitive biases that perpetuate depressive symptoms (Amado-Boccara et al., 1995; Culang et al., 2009; Gao et al., 2022; Joormann & Gotlib, 2010; Shilyansky et al., 2016; Vai et al., 2015). Furthermore, a meta-analysis on the effects of antidepressants on cognitive functioning in MDD found that antidepressants have a modest, positive effect on divided attention, executive function, immediate memory, processing speed, recent memory and sustained attention in MDD patients (Prado et al., 2018).

Cognitive impairments – particularly deficits in executive function and attention – are closely tied to maladaptive thought processes like rumination (Lehmann et al., 2016; Vidal et al., 2020). Rumination, a hallmark of MDD, involves repetitive, self-focused negative thinking, often exacerbating emotional dysregulation and hindering recovery (Brinker & Dozois, 2009; Watkins, 2008). Recent cognitive and neuropsychological research offers additional insights through the impaired disengagement hypothesis, which posits that the persistence of negative thoughts in MDD arises from impaired attentional disengagement from negative self-referent information (Koster et al., 2011). This framework helps bridge psychopathology and cognitive science research, suggesting that deficits in cognitive control and cognitive inflexibility – especially within attention networks – contribute to the inability to regulate negative thought patterns effectively (Connolly et al., 2014; Davis & Nolen-Hoeksema, 2000; van Vugt et al., 2018). Our findings of higher FC between DMN, SN, ECN in antidepressant responders compared to non-responders may, therefore, provide a mechanistic explanation for the impaired executive function, which likely perpetuate rumination and emotional dysregulation. This dysfunctional connectivity pattern may then anchor non-responders in maladaptive cognitive cycles, thereby indirectly contributing to poorer antidepressant efficacy and sustained depressive symptomatology.

Additionally, significant positive correlations between FC and illness severity have been observed within the SN and its interactions with the DMN and SMN. The FC between regions associated with emotional salience detection and self-referential processing (e.g., the SN-DMN axis) and between nodes within the SN and SMN’s significant relation to illness severity underscore how network-level dynamics may reflect the interplay between cognitive function and depressive symptoms (Hamilton et al., 2011; Manoliu et al., 2014; Y. Wang et al., 2022). Specifically, these findings align with aforementioned associations between cognitive impairment and FC, suggesting that the observed correlations between FC and illness severity may represent underlying mechanisms that influence antidepressant response. Effective antidepressant treatment may act by rebalancing connectivity across these networks, thereby alleviating depressive symptoms (Andrade & Rao, 2010; Saberi et al., 2024). This highlights the potential for FC to serve not only as a marker for illness severity but also as an indicator of treatment efficacy, with its modulation potentially influencing the interplay of cognitive function, emotional regulation, and antidepressant responsiveness (Dam et al., 2022; Lantrip et al., 2017).

The present study demonstrated the importance of intrinsic FC patterns in elucidating the neurobiological mechanisms underlying antidepressant response in MDD. Our findings reveal distinct connectivity profiles in antidepressant responders versus non-responders, highlighting enhanced FC within networks responsible for emotion regulation, cognitive control, and attention in responders. These patterns likely reflect mechanisms supporting improved emotional and cognitive flexibility, essential for therapeutic success. Despite these strengths, the cross-sectional design of the study precludes definitive conclusions about the causal relationship between FC changes and treatment response. Longitudinal studies with larger, more diverse samples are required to clarify whether these connectivity patterns represent inherent traits predicting response or are dynamically modulated by antidepressants. Furthermore, as this was not a controlled clinical trial, we were unable to standardize the type of antidepressant used. While most patients received SSRIs, the inclusion of other serotonergic medications introduces potential heterogeneity that could confound the results. Future studies may benefit from sensitivity analyses focusing on homogeneous subgroups, such as those receiving only sertraline, to minimize pharmacological confounds. Lastly, the focus on antidepressant-naïve or washed-out patients limits the generalizability of our findings to individuals with treatment-resistant depression or those on combination therapies. While this analysis focused on individual seed-to-cluster and ROI-to-ROI FCs, future studies may benefit from deriving composite FC biomarkers by averaging hub-level DMN or DMN–CCN connectivity, which have been consistently implicated in antidepressant response. Such approaches may enhance the robustness and clinical utility of FC-based biomarkers and facilitate more accurate prediction of treatment outcomes.

Nevertheless, this study reinforces and extends previous evidence linking DMN, SN, and ECN connectivity to antidepressant efficacy, contributing valuable insights to the field. By emphasizing the role of network-level connectivity in shaping emotional and cognitive outcomes, our findings offer a foundation for advancing personalized treatment strategies. Addressing the outlined limitations in future research will be critical to refining our understanding of these mechanisms. Even so, this study represents a meaningful step toward developing more targeted and effective therapeutic approaches for MDD, underscoring the potential of connectivity-based biomarkers to guide clinical interventions and improve outcomes.

## Data Availability

Due to privacy and ethical restrictions, the clinical and neuroimaging data supporting the findings of this study are not publicly available. However, de-identified data may be made available from the corresponding author upon reasonable request.

## Acknowledgments

We appreciated all participants for taking part in this study.

## Ethical standards

The authors assert that all procedures contributing to this work comply with the ethical standards of the relevant national and institutional committees on human experimentation and the Helsinki Declaration of 1975, as revised in 2008.

